# Intimate Partner Violence Victimization and Perpetration among U.S. Adults during COVID-19: A Brief Report

**DOI:** 10.1101/2020.06.08.20125914

**Authors:** Maxine Davis, Ohad Gilbar, Diana Padilla-Medina

## Abstract

**Importance:** Anecdotal evidence such as increased calls to domestic violence (DV) hotlines across the globe suggest that there may be an increase of IPV prevalence in association with the COVID-19 outbreak; however, no study has investigated this phenomenon empirically.

**Objective:** To evaluate the association between COVID-19 related conditions and recent use or experience of IPV (since the pandemic outbreak in the U.S).

**Design, Setting, and Participants:** This cross-sectional study analyzed data collected online from a sample of noninstitutionalized adults (age 18+) in the U.S. (N=2,045). More than half of the sample self-identified as being in an intimate relationship at the time of the study.

**Main Outcomes and Measures:** A four-item tool was used to assess IPV perpetration and victimization since the outbreak of COVID-19. The rapid tool inquired about two forms of IPV, psychological and physical. Participants self-reported demographic data and recent health histories, including COVID-19 tests results, related symptoms and degree of personal social distancing. We hypothesized that COVID-19 related factors would increase risks of IPV.

**Results:** In this study, self-reported COVID-19 impacted respondents had an increased risk of IPV victimization and perpetration. Among those who reported having symptoms consistent with coronavirus, but were denied access to testing, psychological IPV victimization was 3 times greater than those who did not (Exp[B] =3.15, [1.19, 2.29] p <.05). For participants who reported testing positive to COVID-19, the odds of using psychological IPV (Exp[B] =3.24, [1.18, 8.89] p <.05) and physical IPV (Exp[B]=3.02, [1.12, 8.17] p <.05) against an intimate partner increased by more than 3 times.

**Conclusions and Relevance:** Patient education and community outreach/health care system initiatives focused on IPV risk behaviors may help reduce the potential development of IPV. Continued surveillance is imperative to improve health and well-being along with effective intervention development and implementation.

## INTRODUCTION

As of May 20, 2020, more than 1.5 million cases of coronavirus disease 2019 (COVID-19) were reported in the United States (U.S.)^1^. Patients with underlying comorbidities are excessively affected by COVID-19. ^2,3^ Black/African-Americans, subjects of longtime systemic/institutional/structural racism are most disproportionately hit by the pandemic, along with Hispanic/Latinx populations.^1,2,3^ Referred to as a “shadow” pandemic, intimate partner violence (IPV), a pre-existing public-health problem, has reportedly intensified and is expected to rise amid the crisis.^4^ However, studies examining connections between the COVID-19 outbreak and IPV are skim.^5,6^ Social scientists, medical professionals, and public-health experts have called for urgent research into this area as a critical step in responding to mounting and/or anticipated IPV.^7,8,9,10,11^ This study aimed to characterize risk of IPV in relation to COVID-19.

## METHODS

This was a cross-sectional observational study of adults. Participants were recruited by a survey company (Qualtrics) from an online research panel using stratified quota sampling to ensure sample characteristics of sex, age, and race/ethnicity were representative of the U.S. Inclusion criteria required respondents to: live in USA; be age 18+. Study procedures were approved by a university institutional review board. Informed consent was obtained from participants. For participation, each respondent was distributed points valuing $4.80. Pilot-testing started March 17, 2020; data collection ended May 1, 2020. Majority of participants completed the survey April 22^nd^ - April 30^th^. Around 33750-37500 people were invited or may have seen an invitation; 3750 expressed interest in the study. After eligibility screening, refusals, and removal of poor-quality responses, 2,045 participants’ data were collected.

### Dependent-Variable

IPV was measured using the Jellinek inventory for assessing partner violence (J-IPV). The J-IPV is a 4-item screening tool developed to assess IPV victimization and perpetration in patients entering substance abuse treatment^12^. Drawing on a recent review of brief IPV screening measures^13^, the J-IPV was selected for its strong psychometric properties and brevity.

Participants were asked about recent engagement or experience of IPV, e.g.: “since the coronavirus crisis started, the situation with your partner got so out of hand that you acted in a threatening way to your partner, or threatened to hurt him/her”? ‘Yes’=1; ‘No’=0.

### Independent-Variables

#### COVID-19 status

Participants were asked 8 questions about their personal health, e.g.: “I tested positive to being a coronavirus carrier?” Yes=‘1’; No=‘0’

#### Social distancing restrictions

Participants were asked 7 questions about their personal condition in relationship to social changes/restrictions, e.g.: “I lost my job because of the coronavirus crisis.” Yes=‘1’; No=‘0’

#### COVID-19 cases per state

Based on a COVID-19 index reported by Johns Hopkins University^14^, individual states were coded as a 4-level ordinal variable, ranging low-high and used to indicate respondents’ level of COVID-19 exposure.

### Statistical Analysis

We calculated descriptive statistics to examine distributions of variables and generalized linear modeling (GLM) to examine covariate levels of cases by state. Spearman nonparametric correlation analyses were used to determine whether IPV victimization and perpetration were significantly associated with COVID-19 status and social distancing restrictions. Four binary logistic regressions within a GLM framework were performed to determine impact of coronavirus exposure, government restrictions, and social distancing differences on IPV probability, while controlling for sociodemographics. Covariates evaluated in univariate analysis (*P*□<□.10) were included in multivariable analysis. For dependent variables, missing data were low (0%-3.4%). Analyses were performed using SPSS version-25.

## RESULTS

Mean age of the sample was 46.63 years (SD = 17.19, range = 18–91 years); 49.9% (n = 1020) were ‘female’ and 1.5% (n = 30) of participants were ‘other’. Majority of individuals indicated they were in an intimate relationship (58.2%, n = 1183); 87.4% completed a college-degree; 47.2% of household incomes were $49k or above. Racial identity was: 62.6% White/European-American, 11.9% African-American, 3.3% Hispanic/Latino, 2% Asian-American, 20.1% other. Sexual orientation was: 80.1% heterosexual, 3.8% gay/lesbian, 5.9% bisexual, 5.5% other.

Spearman nonparametric correlation analyses, presented in Table 1, examined items as quoted in the survey. One set of items were positively correlated with IPV, while another set were negatively correlated. We examined group differences by state-level intensity of positive COVID-19 cases in relationship to IPV using GLM (see Table 2). Differences between coronavirus intensity groups were estimated for Psychological Victimization: *χ*_(3)_^2^=9.29, p<.05; Physical Victimization: *χ*_(3)_^2^=10.84, p<.05; and Psychological Perpetration: *χ*_(3)_^2^=9.99, p<.05. We then employed a group mean ranking, based on post-hoc pairwise comparisons between expected violence rates. These comparisons were subject to the Bonferroni correction for multiple comparisons. As expected, group 4 had significantly higher rates of IPV victimization than group 1.

**Table 1.** Personal coronavirus health conditions, restrictions condition, social/occupational changes during the crisis correlated with IPV

|  | Victims of<br>Psychological<br>IPV | Victims of<br>Physical<br>IPV | Perpetration of<br>Psychological<br>IPV | Perpetration of<br>Physical<br>IPV |
| --- | --- | --- | --- | --- |
| I tested positive to being a Corona Virus carrier | .27** | .19** | .26** | .24** |
| I have had symptoms consistent with Corona Virus (e.g., fever, coughing, shortness of breath, etc.). | .13** | .17** | .10** | .12** |
| I have had symptoms consistent with Corona Virus, but I was denied access to testing. | .27** | .19** | .23** | .22** |
| I have had symptoms consistent with Corona Virus, but I did not want to get tested. | .21** | .18** | .21** | .16** |
| I was tested for Corona Virus, but results came back negative. | .24** | .23** | .22** | .19** |
| I had cold or flu-like symptoms and was ordered by a medical professional to stay at home for a period of 14 days to 40 days. | .12** | .11** | .12** | .12** |
| I have not had any symptoms consistent with Corona Virus. | -.30** | -.25** | -.26** | -.22** |
| Nothing has changed since the crisis started. | .26** | .13** | .26** | .20** |
| Due to government restrictions, I am practicing one of the following measures: social distancing, isolation and/or quarantine. | -.16** | -.10** | -.15** | -.10** |
| Due to personal choice, I am practicing one of the following measures: social distancing, isolation and/or quarantine. | -.03 | -.03 | -.02 | -.10 |
| I am not practicing any of these measures. | .15** | .12** | .15** | .12** |
| I'm not going to work because of the situation. | .14** | .01** | .11** | .10** |
| I lost my job because of the coronavirus crisis. | .09* | .12** | .11** | .14** |
| My social life has changed. I tend to avoid social meetings. | .01 | -.03 | -.02 | -.002 |
| I'm more alone. |  |  |  |  |
| My family life has changed. | .01 | -.03 | .02 | .10 |

This table shows Spearman (non-parametric correlations) that were performed across those with any type of intimate relationship (N=1172) of the 8 Covid-19 status, the 7 Social distancing restrictions and IPV.
Note: IPV- intimate partner violence

**Table 2.**
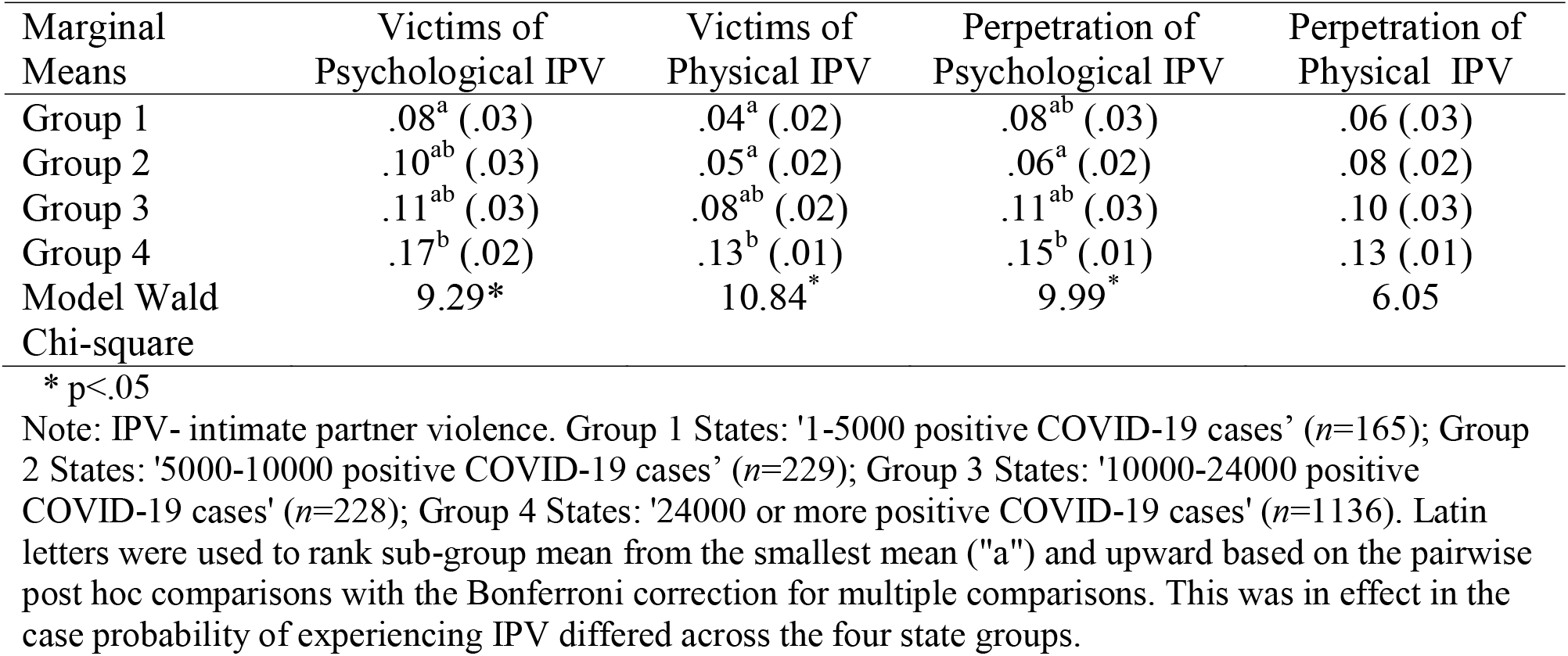
Group differences between state level of coronavirus cases and IPV

IPV usage and experiences were regressed on COVID-19 conditions. Prior to running regression models, we examined independent variables for multicollinearity. Collinearity diagnosis revealed small collinear effects between independent indicators. Tolerance was nearly for each independent indicator; variance inflation factor (VIF) was slightly above 1.00 for all indicators (following recommendations^15^; VIF<10). Table 3 presents variables retained in the models. Among those endorsing “I tested positive to being a Corona Virus carrier”, odds of experiencing psychological IPV increased by more than 3.5 times (Exp [B] =3.77, [1.36, 10.42] p <.05), and experiencing physical IPV increased by more than 2.5 times (Exp[B] = 2.77, [1.04, 7.36] p <.05). Respondents who were tested for the virus but received negative results were twice as likely of being victims of IPV than those not tested (Exp[B] =2.09, [1.23, 3.57] p <.01; Exp[B] =2.20, [1.26, 3.86] p <.01). In three IPV conditions, when respondents had no symptoms, they showed lower probability to experience violence (psychological; Exp[B] =0.59, [.35, .97]p <.05; physical: Exp[B] =0.56, [.32, .96]p <.05) and use psychological violence (perpetration: Exp[B] =0.57, [.33, .96] p <.05). If a respondent reported they were not going to work because of COVID-19, probability of being a victim of psychological IPV increased by 62% (Exp[B] =1.62, [1.04, 2.52] p <.05). For participants reporting job loss as the result of COVID-19, probability of using IPV increased by more than 2.5 times (psychological: Exp[B] =2.56, [1.26, 5.18] p <.01; physical: Exp[B] =3.21, [1.67, 6.58] p <.01). Since COVID-19, men were 2 times more likely to use physical violence in comparison to women (Exp[B] =2.16, [1.31, 3.56] p <.01) and older respondents were less likely to experience or use IPV, compared to younger respondents. State levels of COVID-19 were not found to be a significant predictor of IPV.

**Table 3.**
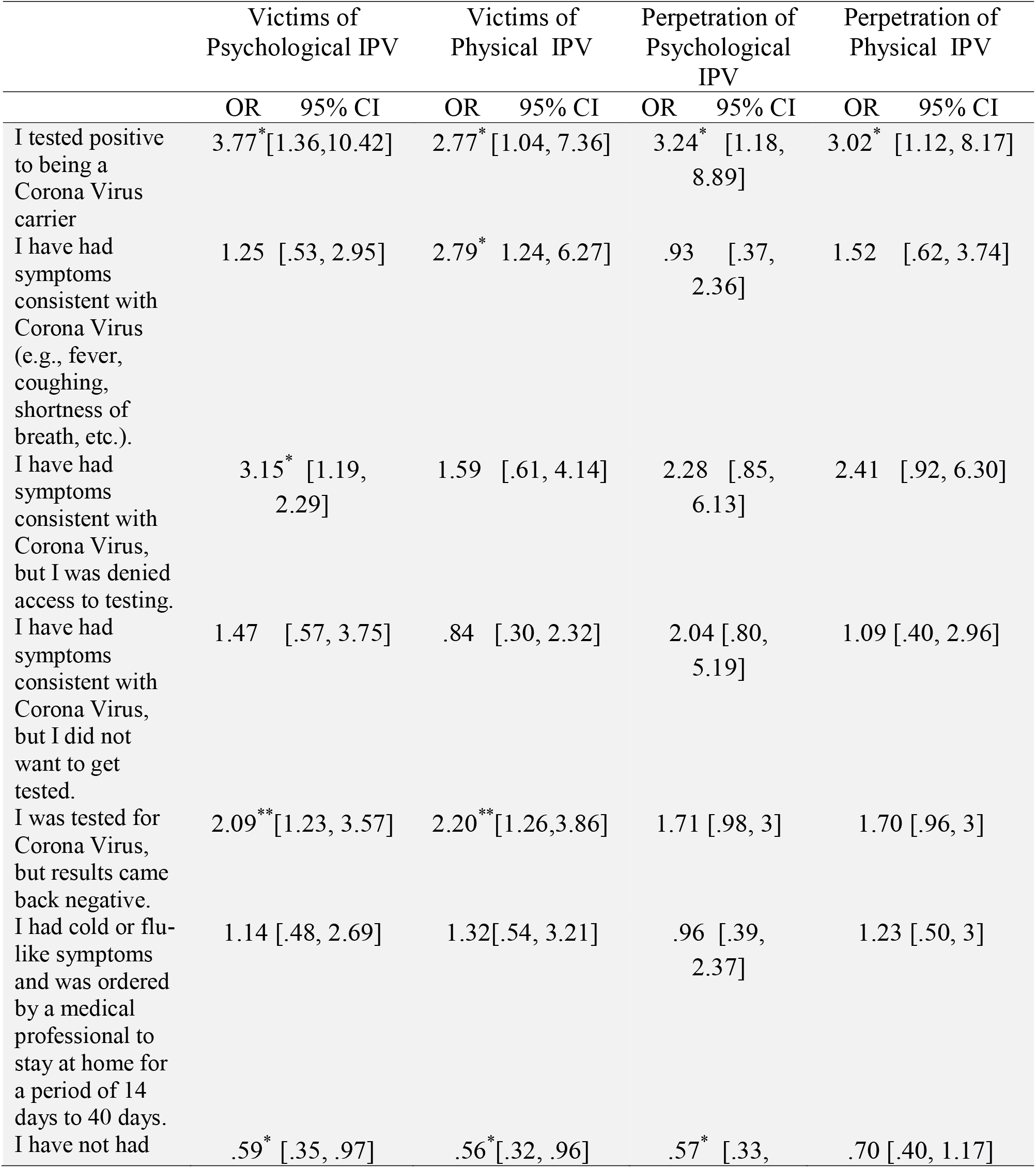
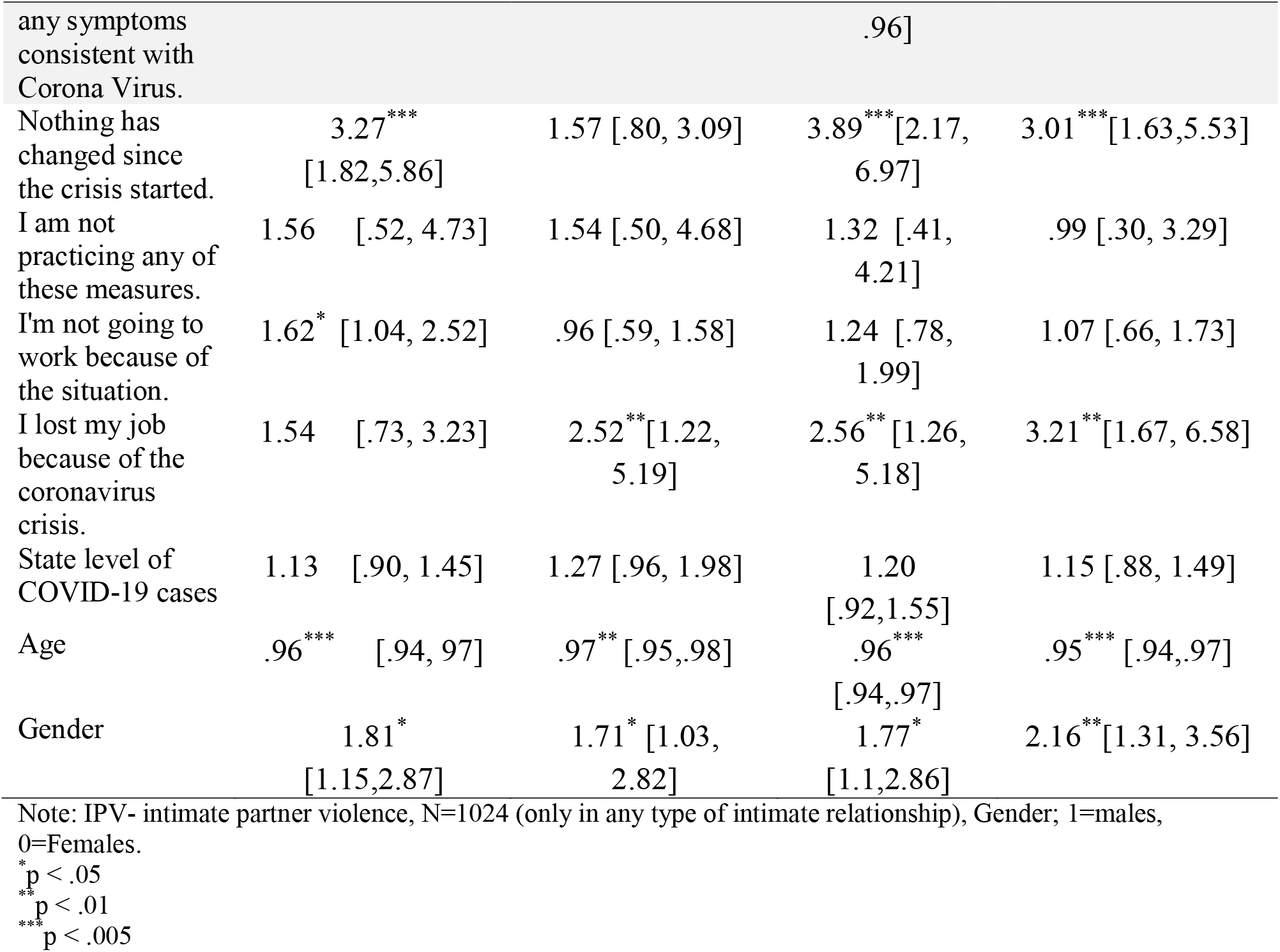
Binary logistic regression results assessing personal coronavirus health conditions, restrictions condition as Predictors of IPV psychological and physical victimization and perpetration:

## DISCUSSION

People reporting COVID-19 positive were nearly 3 times more likely to experience IPV and 3 times more likely to use IPV. Residents in states with low-levels of COVID-19 were impacted with IPV at lower rates than states with higher virus spread. However, state level of COVID-19 outbreak was not found to be a significant IPV risk factor. People who lost their jobs due to COVID-19 were 2.5-3 times more likely to perpetrate IPV and experience physical IPV. The results must be interpreted with caution due to study limitations such as utilizing self-reported data from a cross-sectional survey. An additional limitation is that the models in this brief report did not include key variables such as race/ethnicity. A more comprehensive model is expected to improve explanatory power and yield more precise results. These findings should *not* be interpreted as an endorsement to disengage in social distancing practices. Rather, these results emphasize need for an ongoing public-health response to IPV, including further investigation, funding for development/evaluation of interventions, and implementation of evidence-based practices.

## CONCLUSION

This study provides initial evidence the pandemic is associated with increased likelihood of IPV, which seems to support emerging evidence^16^. The relationship between IPV and COVID-19 has important implications for scientific study, medical, nursing, public-health and social work practice.

## Data Availability

N/A

